# *De novo* variant analysis of childhood-onset obsessive-compulsive disorder in the French-Canadian population

**DOI:** 10.1101/2024.09.17.24313639

**Authors:** Kate Bornais, Jay P. Ross, Zoe Schmilovich, Miranda Medeiros, Dan Spiegelman, Bernard Boileau, Jean-Jacques Marier, Ghislain Laurin, Patrick A. Dion, Guy A. Rouleau

## Abstract

Childhood-onset obsessive-compulsive disorder (OCD) is a neuropsychiatric disorder with a strong genetic component. *De novo* variants (DNVs) have been shown to have a role in childhood-onset OCD, but to date, no DNV analysis has been performed in patients from a genetically isolated population. Here, we aimed to investigate the impact of rare *de novo* single nucleotide variants (dnSNVs) on childhood-onset OCD risk in the French-Canadian population. In a cohort of 36 French-Canadian trios comprised of 36 probands with childhood-onset OCD and 72 unaffected parents, we identified 34 dnSNVs harboured in 34 different genes. We found that four of these genes were previously associated with OCD, replicating their contribution to its risk. We also observed complete overlap between our 34 candidate genes and genes associated with 11 related neuropsychiatric disorders, supporting a shared underlying genetic susceptibility across psychopathologies. Among genes harbouring DNVs across three childhood-onset OCD cohorts, we observed an overrepresentation of genes involved in clathrin-dependent endocytosis (GO:0072583; *p-adj* = 0.0498) and phosphatidylinositol binding (GO:0035091; *p-adj* = 0.0431), offering potential biological mechanisms underlying childhood-onset OCD. No association was found between the number of dnSNVs in childhood-onset OCD probands and OCD symptom severity. Altogether, this study offers a framework for performing DNV analyses of complex disorders in genetically isolated populations. We have provided the first list of candidate childhood-onset OCD genes in the French-Canadian population.

## Introduction

Obsessive-compulsive disorder (OCD) is a common neuropsychiatric disorder with a lifetime prevalence of ~1-3% in the general population^1^. Individuals with OCD experience recurrent intrusive thoughts or sensations (obsessions) and, in response, excessively perform repetitive behaviours (compulsions) aiming to alleviate their anxiety^2^. Symptoms of OCD can cause such substantial distress and/or impairment in crucial areas of functioning that the World Health Organization currently ranks OCD among the top 10 most debilitating disorders of any kind^3,4^.

Childhood-onset OCD represents a clinically distinct patient subgroup of OCD characterized by the manifestation of symptoms before 18 years of age^5^. While the aetiologies of both childhood- and adult-onset OCD are multifaceted and remain poorly understood, studies focused on one subtype allow for a more targeted investigation of the genetic landscape of OCD and can uncover genetic factors contributing to the distinct clinical trajectory of a particular subgroup. Childhood-onset OCD, as compared to adult-onset OCD, is associated with greater global symptom severity and is predicted to have a stronger genetic component, with heritability estimates being ~45-65% for childhood-onset OCD and ~27-45% for adult-onset OCD^1,5,6^. Therefore, childhood-onset OCD cohorts are better suited for genetic investigations.

*De novo* variants (DNVs) are rare genomic alterations that arise in parental germ cells (eggs or sperm) or zygotes (fertilized eggs) but are not present in the genomes of either biological parent^7,8^. DNVs are frequently more deleterious than inherited variants due to their lack of evolutionary selection and typically impart large phenotypic effects^8,9^. As such, DNV approaches have shown great success for systematic risk gene discovery in complex neuropsychiatric phenotypes^9,10^. Given that DNVs are predicted to play a larger role in severe early-onset psychiatric disorders than late-onset disorders, DNV analyses are especially valuable for investigating the genetic basis of childhood-onset psychopathologies^7^.

The role of rare DNVs in risk for childhood-onset OCD is well-established^11,12^. It has been estimated that DNVs contribute to risk in ~22% of childhood-onset OCD cases^11,12^. A significant exome-wide enrichment of damaging DNVs has also been previously observed in cases with childhood-onset OCD relative to their unaffected parents and general population controls^11,12^. Additionally, two high-confidence risk genes – chromodomain-helicase-DNA-binding protein 8 (*CHD8*) and signal peptide, CUB domain and EGF like domain containing 1 (*SCUBE1*) – were also identified in childhood-onset OCD through DNV analyses of whole-exome sequencing (WES) data of parent-child trios^11,12^. However, the number of DNVs in childhood-onset OCD remains limited and many of the associated genes have yet to be replicated.

To date, no DNV study of childhood-onset OCD has been performed in a genetically isolated founder population. These populations are characterized by a limited gene pool and a high degree of shared ancestry. Studying DNVs within genetically isolated populations offers an opportunity to uncover novel genetic contributors to disorders that may have arisen independently within specific groups^13^. Additionally, using genetically homogenous populations for DNV analyses can increase confidence in calling true DNVs. This is due to the phenomenon of “missed heterozygotes”, wherein heterozygous variants can be wrongly sequenced as absent from a parent due to technical or computational errors, leading to apparent DNVs in children that were actually inherited^14,15^.

The French-Canadian population of Québec, Canada, is a widely recognized genetic isolate that has yielded many findings that other large-scale studies would have missed due to population background heterogeneity^16^. The historical founder effect of the French-Canadian population and their subsequent isolation due to linguistic, religious, and geographic barriers in the 17^th^ century resulted in a distinct and homogenous genetic background that requires its own complete catalogue of disorder-associated variants within the population^16^. The French-Canadian population also often allows access to familial data, typically with data available from both parents, and are deeply clinically phenotyped. Considering OCD is one of the most prevalent psychiatric disorders in Québec, with a provincial prevalence of ~1.2%^17^, a DNV analysis of French-Canadian cases with childhood-onset OCD represents an important avenue of investigation.

Here, we performed a DNV analysis of childhood-onset OCD probands and their unaffected biological parents from the French-Canadian population to identify genetic factors associated with childhood-onset OCD risk and symptom severity within the population. In doing so, we have developed a framework for conducting DNV analyses of complex psychiatric disorders in genetically isolated populations.

## Methods

The methodological framework presented below can be visualized in Figure 1.

**Figure 1.**
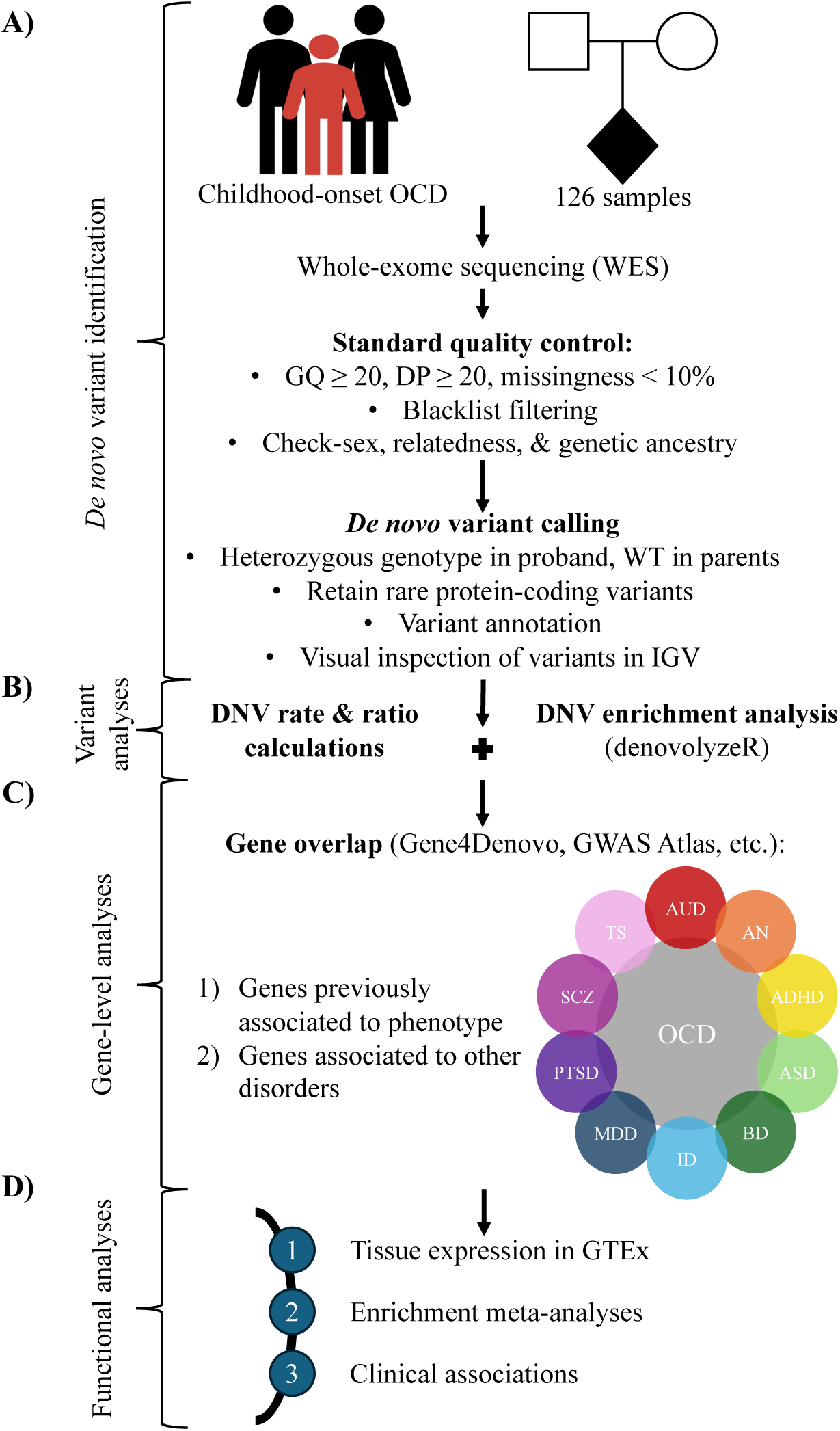
A framework for performing DNV analyses of complex psychiatric disorders in genetically isolated populations. A) The discovery flow of DNVs from trios, including sample collection, standard data QC, DNV calling and annotation, and visual inspection of DNVs. B) Variant-level analyses, including *de novo* mutation rate and ratio calculations and enrichment analyses of variant classes. C) Gene-level analyses, including examining for overlap within and between phenotypes using publicly available databases. D) Functional analyses, including tissue expression and enrichment analyses of candidate genes as well as correlational analyses of DNVs with clinical data.

### Cohort overview

In total, 42 complete trios consisting of 42 probands with childhood-onset OCD and 84 unaffected parents from Québec, Canada, were included in this study. Initial inclusion criteria required all three trio participants to have self-reported as French Canadian. We excluded 6 trios where one or more members self-reported as another ancestry. Samples underwent WES at Axeq Technologies Psomagen Laboratory (Rockville, Maryland) with the SureSelect XT Human All Exon enrichment kit version 7 (Agilent) using the Illumina NovaSeq6000 S4 Sequencing System and following standard protocols. Reads were aligned to the *Homo sapiens* (human) reference genome build GRCh37 (hg19). All OCD diagnoses were made by psychiatrists at the Centre Hospitalier Universitaire Sainte-Justine (Montréal, Québec, Canada) and were based on the Diagnostic and Statistical Manual of Mental Disorders, fourth edition (DSM 4) criteria^18^. Additional inclusion criteria required probands to have received their OCD diagnosis before 18 years of age and have no known history of OCD in first-degree relatives. Sample details are summarized in Supplementary Table 1.

### Quality control

Variant- and sample-level quality control (QC) were performed using the Genome Analysis Toolkit (GATK) version 3.8 (Supplementary Table 2)^19^. Variants were filtered according to the following standard QC thresholds: genotype quality (GQ) ≥ 20, depth of coverage (DP) ≥ 20, and missingness < 10%^11,12^. Variants in blacklist regions of the genome were also removed^20^. Biological sex was imputed for each sample to ensure proper sample identity using the PLINK2.0 “check-sex” function^21^. A kinship analysis was then conducted using the KING algorithm to ensure that trios displayed biological parent-child relationships, parents were in non-consanguineous relationships, and families were unrelated^22^. Principal component analysis (PCA) was performed to cluster individuals by genetic ancestry and account for population stratification^23^. WES data of each participant were compared to the 1000 Genomes reference dataset to filter out samples that did not cluster with the expected European genetic ancestry and to ensure tight clustering indicative of a genetically homogenous study population^24^.

### *De novo* variant calling, annotation, and filtration

The initial set of DNVs was filtered for heterozygous genotype in the proband and wildtype in both parents using the GATK version 3.8^19^, and then annotated using the Ensembl Variant Effect Predictor (VEP)^25^. DNV calls were included in the downstream analyses if they were protein-coding, observed only once in the cohort, and had a minor allele frequency (MAF) < 1% in the non-Finnish European reference population of the Genome Aggregation Database (gnomAD)^9,11,12,26^. Missense, nonsense, splice site, and synonymous single nucleotide variants (SNVs) were retained. All *de novo* SNV (dnSNV) calls passing these criteria were manually confirmed through visual inspection in the Integrative Genomics Viewer (IGV). Any call detected in < 30% of proband reads or > 5% of parent reads at the call site were removed^12^. Variants that displayed strong strand biases and/or false positives due to misalignment of reads or low base quality (Phred Quality Score (Q) < 20) were also removed^27^. Probands with more than 5 dnSNV calls that met these criteria were excluded from the analyses^11,12^. The deleteriousness of the final set of dnSNVs was determined based on their Combined Annotation Dependent Depletion (CADD) score^28^.

### *De novo* mutation rate and probability analyses

We calculated the average number of dnSNVs per proband, from which we estimated the observed *de novo* mutation rate of the French-Canadian childhood-onset OCD probands. The observed *de novo* mutation rate was calculated as follows:

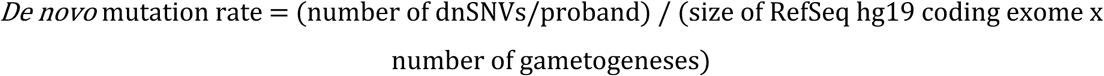

where the size of the *Homo sapiens* (human) reference genome (RefSeq) GRCh37 (hg19) coding exome was 33,828,798 base pairs (bps)^29^ and the number of gametogeneses was two (spermatogenesis and oogenesis). We then calculated the average number of non-synonymous and synonymous dnSNVs per proband to determine the observed ratio of non-synonymous to synonymous dnSNVs. The mean number of non-synonymous and synonymous dnSNVs detected in the probands was then compared using a two-tailed paired samples t-test using the *t.test* function from the base R package.

To assess whether there was a genome-wide excess of dnSNVs in different functional variant classes, we used the denovolyzeByClass function from the denovolyzeR R package^30^. Briefly, denovolyzeR implements a mutational model that estimates the probability of a dnSNV occurring in a single copy of each human gene in one generation based on the local sequencing context, from which it can calculate the expected number of dnSNVs for a given population size^30^. The denovolyzeByClass function tabulates the dnSNV probabilities for each variant class (e.g., nonsense, missense, splice site, synonymous, etc.). It then determines whether there is an overrepresentation of dnSNVs in a particular variant class within a population of interest by comparing the expected dnSNV frequencies to those observed using a Poisson framework^30^. For each variant class, the function returns the expected number of dnSNVs, the enrichment ratio (= observed/expected), and *p* values obtained from the Poisson regression^30^.

### Gene-level analyses

Genes harbouring dnSNVs that passed all QC criteria were gathered to generate a list of candidate childhood-onset OCD genes. The literature was mined to identify genes that have been previously associated with OCD through other DNV analyses or genome-wide association studies (GWASs)^11,12,31^. We then compared these genes with our list of candidate childhood-onset OCD genes to identify overlap. Additionally, due to the known clinical and genetic overlap across psychiatric disorders, we assessed whether our candidate childhood-onset OCD genes were associated with other related psychiatric disorders. We compared our candidate gene list to genes previously associated with 11 other psychiatric disorders through DNV analyses listed in the Gene4Denovo database and GWASs listed in the GWAS Atlas, respectively^32,33^. To confirm that our candidate genes were expressed in brain tissues, we input our candidate gene list into the Expression feature of the Genotype-Tissue Expression (GTEx) Project Portal^34^. Using the Multi Gene Query of this feature, we performed a multi-gene search for expression across all available brain tissues^34^. This output a heatmap representing normalized transcript per million (TPM) expression values from RNA-seq data for each gene in the selected tissues^34^.

### Functional enrichment meta-analyses

Genes harbouring DNVs in our French-Canadian childhood-onset OCD probands (*n* = 34 genes) were combined with genes harbouring DNVs in 771 previously reported OCD trios and 41 previously reported OCD quartets (*n* = 871 genes)^11,12^. The combined list of candidate childhood-onset OCD genes (*n* = 905 genes) was input into EnrichR and g:Profiler g:GOSt for functional enrichment meta-analyses^35,36^. Results from the two sources were compared and only common pathways identified by both models to be significantly enriched after false discovery rate (FDR) correction were considered.

### Statistical analyses

Biological sex, age at OCD diagnosis, and the first five ancestry principal components (PCs) were included as covariates in all three regression models below. All *p* values were corrected for multiple comparisons using the Benjamini–Hochberg FDR method available in the *p.adjust* function from the base R package.

Two linear regression models were used to assess the relationship between the number of dnSNVs in probands and parental ages at conception of the probands:

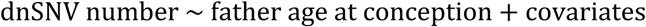

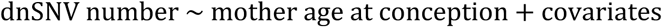

A logistic regression model was then used to examine the effect of the number of dnSNVs in childhood-onset OCD probands on whether OCD symptoms were rated as severe or mild to moderate:

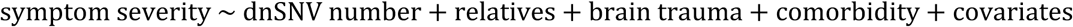

The severity of obsessive and compulsive symptoms was measured using the Children’s Yale-Brown Obsessive Compulsive Scale (CY-BOCS) for 30 out of the 36 included probands at the time of their diagnoses^37^. Total CY-BOCS scores were calculated as the sum of the obsessions and compulsions subscale severity scores^37^. Probands with CY-BOCS scores ≥ 24 were classified as having severe OCD symptoms, while probands with CY-BOCS scores < 24 were classified as having mild to moderate OCD symptoms^37^. The number of first-degree relatives of each proband with a psychiatric diagnosis was included in the model to control for inherited genetic risk factors; history of traumatic brain injury was included in the model to control for a known environmental risk factor in childhood-onset OCD; and presence of a comorbid psychiatric disorder was included in the model to control for the general liability to psychopathologies.

## Results

Following QC, 36 trios (36 probands and 72 unaffected parents) were included in the analyses. Trio probands were 53% female with a mean age at diagnosis of 13.0 ± 2.48 years.

### dnSNVs detected in French-Canadian childhood-onset OCD probands

Our framework directly identified 34 dnSNVs in 34 genes across the 36 French-Canadian childhood-onset OCD probands (0.94 dnSNVs/proband, average CADD score = 16.1) that remained after QC (Table 1). 17 of the 34 (50.0%) dnSNVs were missense variants, three (8.8%) were splice site variants, and 14 (41.2%) were synonymous variants (Table 1). 13 of the 34 (38.2%) dnSNVs were variants ranking in the top 1% of deleterious variants in the human genome (i.e., had a CADD score > 20) (Table 1; Supplementary Figure 1)^28^. The estimated *de novo* mutation rate in our French-Canadian childhood-onset OCD probands was 1.40 x 10^-8^ per nucleotide per generation. The observed ratio of non-synonymous to synonymous dnSNVs was 1.43:1 (0.56 non-synonymous dnSNVs/proband, 0.39 synonymous dnSNVs/proband). We observed no difference between the mean number of non-synonymous dnSNVs and the mean number of synonymous dnSNVs in our childhood-onset OCD probands (t = 1.06, *p* = 0.295). All 34 of the candidate childhood-onset OCD genes were predicted to be expressed in one or more brain tissues (Supplementary Figure 2)^34^.

**Table 1.**
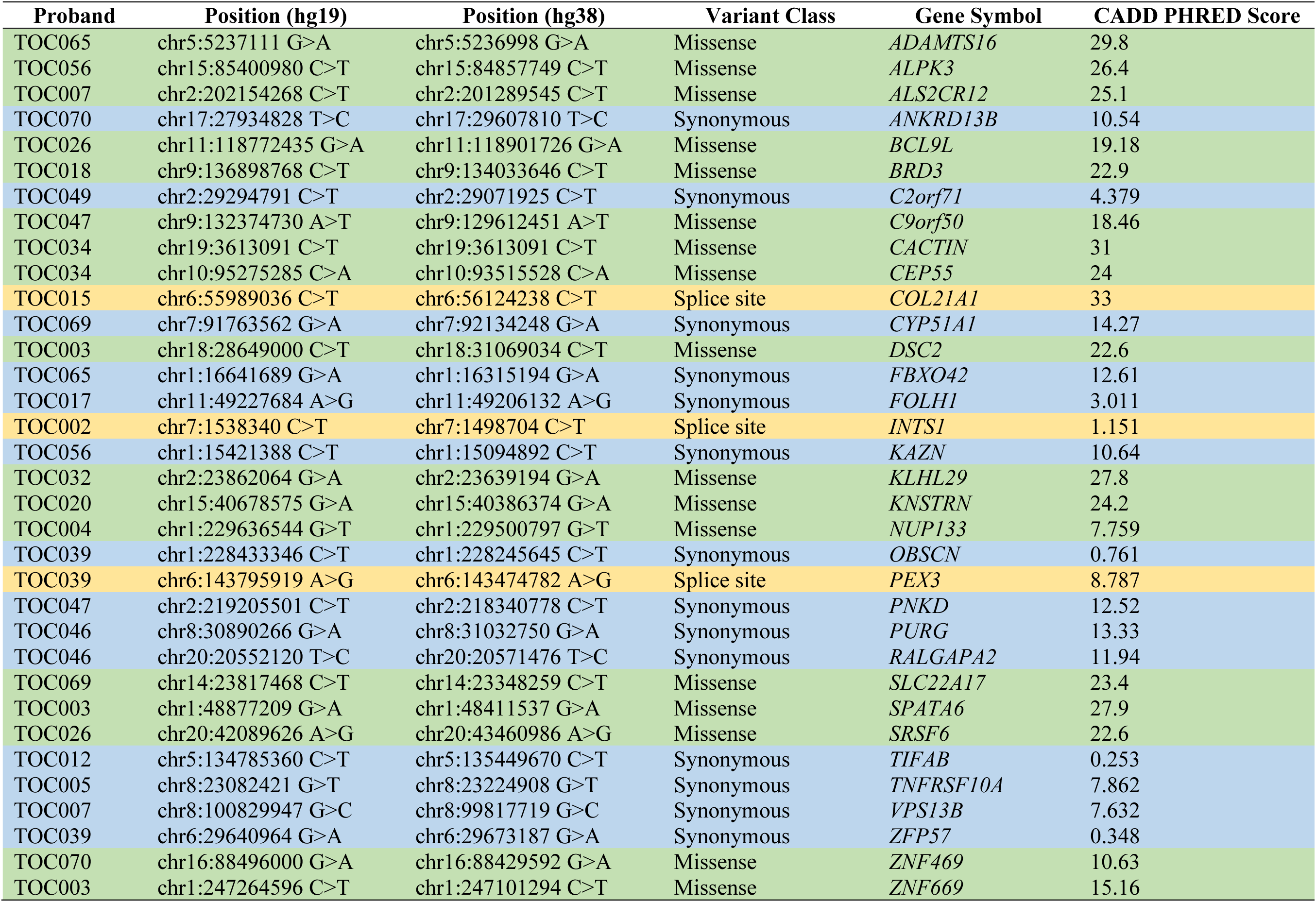
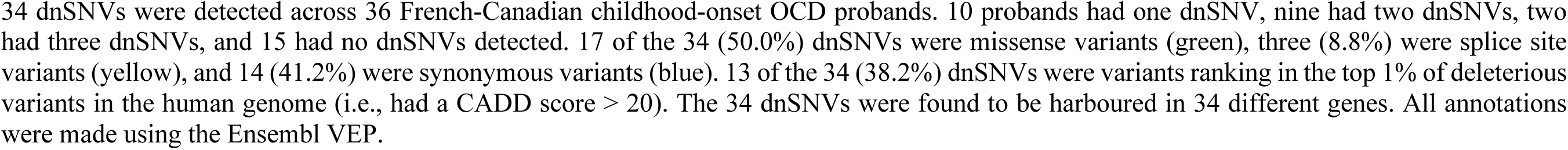
*De novo* single nucleotide variants in French-Canadian childhood-onset OCD probands.

### Evidence for a genome-wide excess of splice site dnSNVs in French-Canadian childhood-onset OCD probands

Using denovolyzeR, we observed no enrichment (i.e., no more observed dnSNVs than expected) of missense dnSNVs (*p* = 0.908, FDR = 0.908) or synonymous dnSNVs (*p* = 0.143, FDR = 0.215) in our probands. However, we found a nominally significant enrichment (i.e., more observed DNVs than expected) of splice site dnSNVs (*p* = 0.0173, FDR = 0.0519).

### Four genes harbouring dnSNVs in French-Canadian childhood-onset OCD probands overlapped with genes previously associated to OCD

To increase confidence in the role of our candidate genes in childhood-onset OCD, we sought to identify repeated observations of DNVs in genes identified to harbour dnSNVs in our French-Canadian probands. Of our 34 candidate genes, two (5.9%) – integrator complex subunit 1 (*INTS1*) and TRAF-interacting protein with FHA domain-containing protein B (*TIFAB*) – were identified in another DNV analysis of WES data of childhood-onset OCD trios^12^. Additionally, two (5.9%) – spermatogenesis associated 6 (*SPATA6*) and zinc finger protein 669 (*ZNF669*) – overlapped with loci from the 2018 OCD GWAS (Table 2)^31^.

**Table 2.**
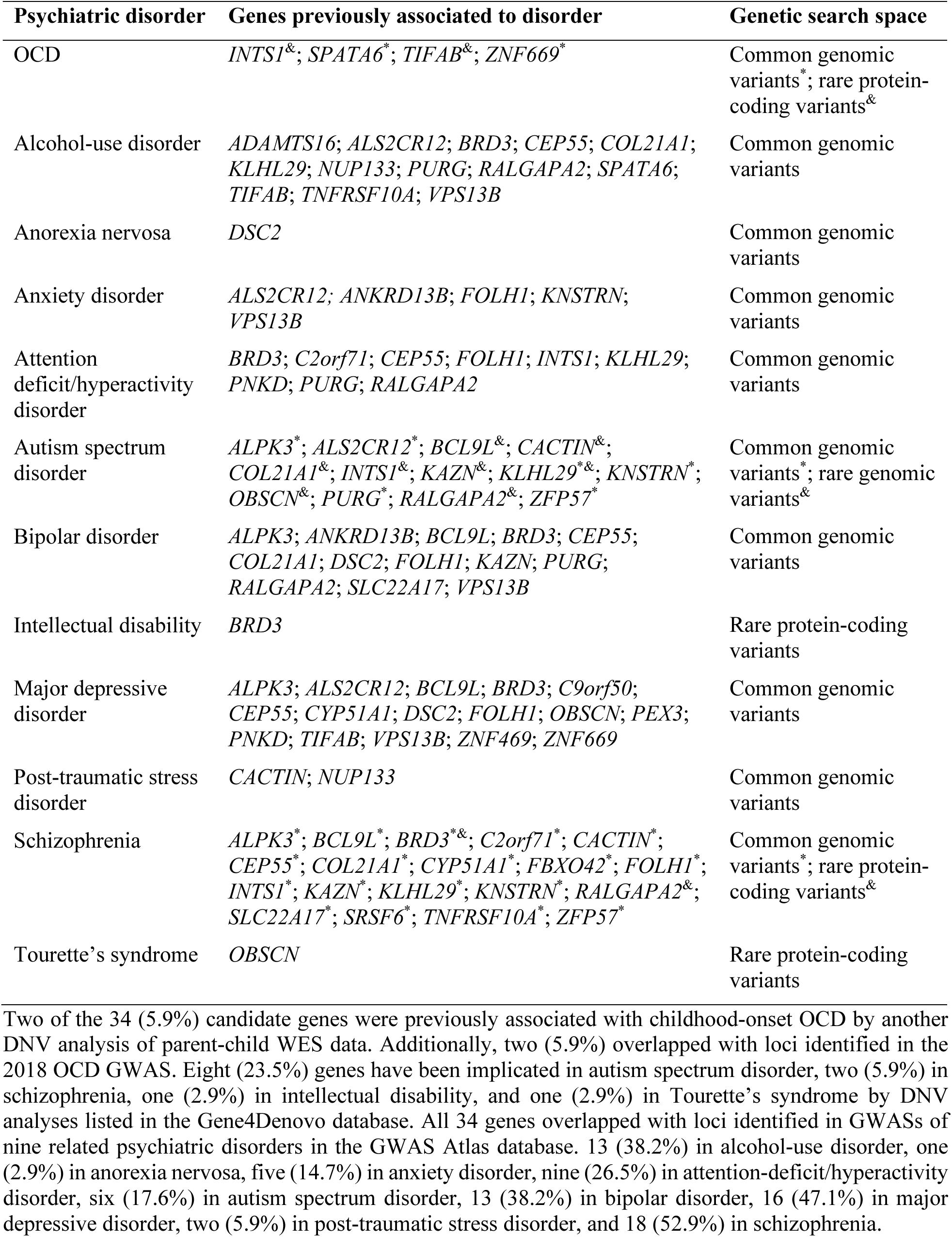
Genes harbouring dnSNVs in French-Canadian childhood-onset OCD probands with previously demonstrated associations to OCD and 11 related psychiatric disorders.

| Psychiatric disorder | Genes previously associated to disorder | Genetic search space |
| --- | --- | --- |
| OCD | *INTS1*<sup>&</sup>; *SPATA6*<sup>*</sup>; *TIFAB*<sup>&</sup>; *ZNF669*<sup>*</sup> | Common genomic variants<sup>*</sup>; rare protein-coding variants<sup>&</sup> |
| Alcohol-use disorder | *ADAMTS16*; *ALS2CR12*; *BRD3*; *CEP55*; *COL21A1*; *KLHL29*; *NUP133*; *PURG*; *RALGAPA2*; *SPATA6*; *TIFAB*; *TNFRSF10A*; *VPS13B* | Common genomic variants |
| Anorexia nervosa | *DSC2* | Common genomic variants |
| Anxiety disorder | *ALS2CR12*; *ANKRD13B*; *FOLH1*; *KNSTRN*; *VPS13B* | Common genomic variants |
| Attention deficit/hyperactivity disorder | *BRD3*; *C2orf71*; *CEP55*; *FOLH1*; *INTS1*; *KLHL29*; *PNKD*; *PURG*; *RALGAPA2* | Common genomic variants |
| Autism spectrum disorder | *ALPK3*<sup>*</sup>; *ALS2CR12*<sup>*</sup>; *BCL9L*<sup>&</sup>; *CACTIN*<sup>&</sup>; *COL21A1*<sup>&</sup>; *INTS1*<sup>&</sup>; *KAZN*<sup>&</sup>; *KLHL29*<sup>&</sup>; *KNSTRN*<sup>*</sup>; *OBSCN*<sup>&</sup>; *PURG*<sup>*</sup>; *RALGAPA2*<sup>&</sup>; *ZFP57*<sup>*</sup> | Common genomic variants<sup>*</sup>; rare genomic variants<sup>&</sup> |
| Bipolar disorder | *ALPK3*; *ANKRD13B*; *BCL9L*; *BRD3*; *CEP55*; *COL21A1*; *DSC2*; *FOLH1*; *KAZN*; *PURG*; *RALGAPA2*; *SLC22A17*; *VPS13B* | Common genomic variants |
| Intellectual disability | *BRD3* | Rare protein-coding variants |
| Major depressive disorder | *ALPK3*; *ALS2CR12*; *BCL9L*; *BRD3*; *C9orf50*; *CEP55*; *CYP51A1*; *DSC2*; *FOLH1*; *OBSCN*; *PEX3*; *PNKD*; *TIFAB*; *VPS13B*; *ZNF469*; *ZNF669* | Common genomic variants |
| Post-traumatic stress disorder | *CACTIN*; *NUP133* | Common genomic variants |
| Schizophrenia | *ALPK3*<sup>*</sup>; *BCL9L*<sup>*</sup>; *BRD3*<sup>*&</sup>; *C2orf71*<sup>*</sup>; *CACTIN*<sup>*</sup>; *CEP55*<sup>*</sup>; *COL21A1*<sup>*</sup>; *CYP51A1*<sup>*</sup>; *FBXO42*<sup>*</sup>; *FOLH1*<sup>*</sup>; *INTS1*<sup>*</sup>; *KAZN*<sup>*</sup>; *KLHL29*<sup>*</sup>; *KNSTRN*<sup>*</sup>; *RALGAPA2*<sup>&</sup>; *SLC22A17*<sup>*</sup>; *SRSF6*<sup>*</sup>; *TNFRSF10A*<sup>*</sup>; *ZFP57*<sup>*</sup> | Common genomic variants<sup>*</sup>; rare protein-coding variants<sup>&</sup> |
| Tourette's syndrome | *OBSCN* | Rare protein-coding variants |

Two of the 34 (5.9%) candidate genes were previously associated with childhood-onset OCD by another DNV analysis of parent-child WES data. Additionally, two (5.9%) overlapped with loci identified in the 2018 OCD GWAS. Eight (23.5%) genes have been implicated in autism spectrum disorder, two (5.9%) in schizophrenia, one (2.9%) in intellectual disability, and one (2.9%) in Tourette's syndrome by DNV analyses listed in the Gene4Denovo database. All 34 genes overlapped with loci identified in GWASs of nine related psychiatric disorders in the GWAS Atlas database. 13 (38.2%) in alcohol-use disorder, one (2.9%) in anorexia nervosa, five (14.7%) in anxiety disorder, nine (26.5%) in attention-deficit/hyperactivity disorder, six (17.6%) in autism spectrum disorder, 13 (38.2%) in bipolar disorder, 16 (47.1%) in major depressive disorder, two (5.9%) in post-traumatic stress disorder, and 18 (52.9%) in schizophrenia.

### Genes harbouring dnSNVs in French-Canadian childhood-onset OCD probands display complete overlap with genes associated with other psychiatric disorders

No OCD-specific gene was identified as all 34 of our candidate genes overlapped with genes previously associated with one or more related psychiatric disorders. 10 of our 34 candidate genes were previously implicated in four related psychiatric disorders (autism spectrum disorder, intellectual disability, schizophrenia, and Tourette’s syndrome) by other DNV analyses (Table 2)^32^. Additionally, all 34 genes overlapped with loci found in GWAS of nine related psychiatric disorders (alcohol-use disorder, anorexia nervosa, anxiety disorder, attention-deficit/hyperactivity disorder, autism spectrum disorder, bipolar disorder, major depressive disorder, post-traumatic stress disorder, and schizophrenia) (Table 2)^33^.

### Functional enrichment meta-analyses of genes harbouring DNVs in childhood-onset OCD probands

Within the combined candidate gene list, we observed that the clathrin-dependent endocytosis Gene Ontology (GO) biological process term (GO:0072583) was significantly overrepresented with eight genes related to that pathway harbouring DNVs across the three studies (*p-adj* = 0.0498) (Figure 2A)^38^. We also observed that the phosphatidylinositol binding GO molecular function term (GO:0035091) was significantly overrepresented with 15 genes related to that pathway harbouring DNVs across the three studies (*p-adj* = 0.0431) (Figure 2B)^38^. The combined list of candidate childhood-onset OCD genes also displayed a significant enrichment of genes expressed in various brain tissues and cells^39^. There was a significant overrepresentation of genes expressed in the spinal cord (*p-adj* = 0.000869); fetal brain (*p-adj* = 0.00267); motor neurons (*p-adj* = 0.00511); cingulate gyrus (*p-adj* = 0.00649); superior frontal gyrus (*p- adj* = 0.0106); astrocytes (*p-adj* = 0.0276); oligodendrocytes (*p-adj* = 0.0411); and prefrontal cortex (*p-adj* = 0.0411) (Figure 2C)^39^.

**Figure 2.**
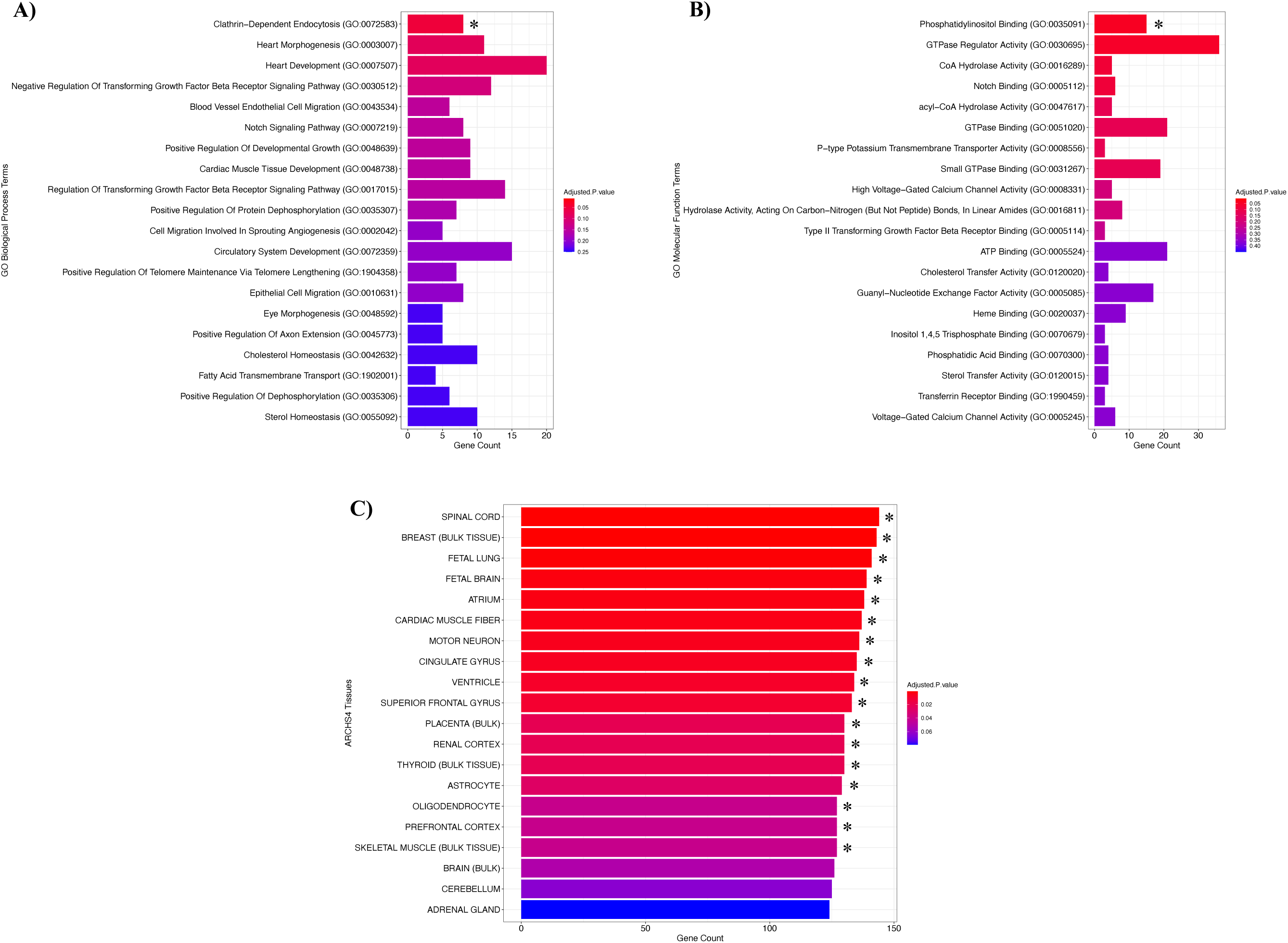
Functional enrichment meta-analyses of genes harbouring DNVs in childhood-onset OCD probands using EnrichR. The input candidate gene list included the 34 genes harbouring DNVs in the present study as well as 871 genes harbouring DNVs in two previous childhood-onset OCD publications (Cappi *et al*. (2020); Halvorsen *et al*. (2021)). All *p* values were adjusted for multiple comparisons using the Benjamini– Hochberg FDR method. * denotes a significant association after FDR correction (*p-adj* < 0.05). Significant pathway enrichments were confirmed in g:Profiler g:GOSt. A) Enrichment of GO biological processes terms. B) Enrichment of GO molecular function terms. C) Enrichment of All RNA-seq and ChIP-seq Sample and Signature Search (ARCHS^4^) Tissue terms.

### Relationship between dnSNVs, parental age, and childhood-onset OCD symptom severity

We modelled the effect of parental ages at proband conception on the number of dnSNVs in our probands using two linear regression models. No association was found between the number of dnSNVs in probands and paternal age at proband conception (*p-adj* = 0.877) or maternal age at proband conception (*p-adj* = 0.761) (Table 3A). Next, we modelled the effect of the number of dnSNVs in the French-Canadian childhood-onset OCD probands on whether OCD symptoms were rated as severe or mild to moderate using logistic regression. No significant association was found between the number of dnSNVs in probands and OCD symptom severity (*p-adj* = 0.336) (Table 3B). Additionally, there was no association identified between the number of first-degree relatives of probands with a psychiatric diagnosis (*p-adj* = 0.336), history of traumatic brain injury (*p-adj* = 0.457), or presence of psychiatric comorbidities (*p-adj* = 0.336) and OCD symptom severity (Table 3B).

**Table 3.** Relationship between the number of dnSNVs in childhood-onset OCD probands and clinical outcomes.

A. Modeling the effects of parental ages at conception on the number of dnSNVs observed in probands.
| Predictor | Paternal age at conception |  |  | Maternal age at conception |  |  |
| --- | --- | --- | --- | --- | --- | --- |
|  | Est. [95%CI] | SE | *p-adj* | Est. [95%CI] | SE | *p-adj* |
| Parental age | -0.0408 [-0.115, 0.0334] | 0.0379 | 0.877 | -0.0671 [-0.186, 0.0467] | 0.0562 | 0.736 |
| Sex | 0.0584 [-0.721, 0.838] | 0.398 | 0.927 | 0.193 [-0.660, 0.906] | 0.378 | 0.917 |
| Age | -0.00736 [-0.163, 0.148] | 0.0793 | 0.927 | 0.0230 [-0.146, 0.176] | 0.0779 | 0.917 |
| PC1 | 0.521 [-1.57, 2.61] | 1.07 | 0.927 | 0.744 [-1.31, 3.05] | 1.05 | 0.917 |
| PC2 | 0.152 [-1.89, 2.20] | 1.04 | 0.927 | 0.236 [-1.86, 2.26] | 0.996 | 0.917 |
| PC3 | -0.438 [-2.50, 1.62] | 1.05 | 0.927 | -0.0640 [-2.26, 2.04] | 1.04 | 0.951 |
| PC4 | 0.607 [-1.42, 2.64] | 1.04 | 0.927 | 0.418 [-1.31, 2.78] | 0.988 | 0.917 |
| PC5 | -1.44 [-3.48, 0.590] | 1.04 | 0.799 | -1.39 [-3.55, 0.523] | 0.984 | 0.736 |

B. Modeling the effect of the number of dnSNVs observed in probands on OCD symptom severity.
| Predictor | Est. [95%CI] | SE | *p-adj* |
| --- | --- | --- | --- |
| dnSNV number | -4.82 [-18.8, -1.11] | 3.40 | 0.336 |
| First-degree relatives | 4.97 [0.829, 17.1] | 3.19 | 0.336 |
| Brain trauma | 2.61 [-2.64, 10.6] | 2.98 | 0.457 |
| Comorbidity | 5.34 [-0.0856, 20.2] | 4.13 | 0.336 |
| Sex | -4.02 [-15.2, 0.488] | 3.38 | 0.352 |
| Age | 1.37 [0.203, 4.57] | 0.974 | 0.336 |
| PC1 | -24.5 [-97.1, -3.60] | 18.5 | 0.336 |
| PC2 | -1.80 [-12.1, 6.77] | 4.38 | 0.681 |
| PC3 | 8.69 [-5.22, 33.3] | 8.92 | 0.450 |
| PC4 | 6.39 [-0.299, 19.3] | 4.21 | 0.336 |
| PC5 | -7.15 [-37.1, 6.82] | 9.03 | 0.468 |

No significant association ( $p < 0.05$ ) before or after FDR correction was detected in any of the three models.
**A.** Two linear regressions modeling the effect of parental age at conception on the number of dnSNVs in childhood-onset OCD probands. 4 probands without paternal birthdate information were excluded from the first linear regression. 5 probands without maternal birthdate information were excluded from the second linear regression. **B.** A logistic regression modeling the effect of the number of dnSNVs in probands on OCD symptom severity measured using CY-BOCS scores. 6 probands without CY-BOCS data were excluded from the analyses.

## Discussion

This is the first DNV analysis of childhood-onset OCD in the French-Canadian population of Québec, Canada. This study provides the first estimate of the *de novo* mutation rate as well as the ratio of non-synonymous to synonymous dnSNVs in French Canadians with childhood-onset OCD. The estimated *de novo* mutation rate of 1.40 x 10^-8^ per nucleotide per generation for our cohort falls within the expected dnSNV rate range of 1.0 – 1.8 x 10^-8^ per nucleotide per generation^7^. This was not surprising as cases of childhood-onset OCD are not expected to have a higher rate of dnSNVs compared to unaffected controls per se. Rather, it is expected that the dnSNVs observed in childhood-onset OCD probands will be more deleterious than those observed in unaffected individuals. The observed ratio of non-synonymous dnSNVs to synonymous dnSNVs of 1.43:1 in our probands is slightly lower than the expected ratio of 2.2:1 reported by previous studies^40^. We propose two explanations for this observation. Firstly, it is possible that this lower-than-expected ratio occurred by chance due to our small sample size. Alternatively, it could point to synonymous dnSNVs as an important source of variation in childhood-onset OCD risk. Future studies will be needed to elucidate the role of synonymous dnSNVs in childhood-onset OCD.

Using our framework, we identified 34 genes that harboured potential childhood-onset OCD-associated dnSNVs specific to the French-Canadian population. The individual expression of all 34 candidate genes in various brain tissues of GTEx samples provides further confidence for their potential roles in childhood-onset OCD risk^34^. The four genes (i.e., *INTS1*, *TIFAB*, *SPATA6*, and *ZNF669*) previously associated to OCD^12,31^ are the most promising candidate genes of childhood-onset OCD identified in this study due to those repeated observations in unrelated probands. The other 30 genes potentially represent novel childhood-onset OCD candidate genes to be screened in OCD cohorts of various origins. The 13 genes harbouring dnSNVs with CADD scores > 20 also represent good candidates for follow-up investigation in childhood-onset OCD due to the predicted severity of these variants^28^.

We did not identify any dnSNVs in genes that were uniquely associated with childhood-onset OCD. This finding supports an overlapping genetic architecture between childhood-onset OCD and 11 related psychiatric disorders (alcohol-use disorder; anorexia nervosa; anxiety disorder; attention deficit/hyperactivity disorder; autism spectrum disorder; bipolar disorder; intellectual disability; major depressive disorder; post-traumatic stress disorder; schizophrenia; and Tourette’s syndrome)^41^. This implies a potential pleiotropic effect and supports the hypothesis of a shared underlying genetic susceptibility across psychiatric disorders and is in line with previous studies which were also unable to identify any OCD-specific genes^41^.

We observed a nominally significant genome-wide excess of splice site dnSNVs within our cohort, suggesting alternative splicing may be an important mechanism in the pathogenesis of childhood-onset OCD. Interestingly, the two high-confidence OCD risk genes identified by previous DNV analyses, *CHD8* and *SCUBE1*, each harboured a splice site dnSNV predicted to decrease splicing efficiency in unrelated childhood-onset OCD probands^11,12^. These results support our ongoing hypothesis of alternative splicing as a potential risk factor in childhood-onset OCD. While future analyses will be needed to confirm and clarify the role of splice site dnSNVs in childhood-onset OCD risk, this study provides the first evidence of an enrichment of rare splice site variants in childhood-onset OCD probands.

Within the combined candidate gene list^11,12^, genes involved in clathrin-dependent endocytosis (GO:0072583) and phosphatidylinositol binding (GO:0035091) were enriched^35,36,38^. Clathrin-dependent endocytosis is a fundamental cellular process whereby cargo molecules undergo internalization from the cell surface and are subsequently transported into vesicles within the cell^42^. It is a particularly essential pathway for proper neural functions^42^. Aberrant clathrin-dependent processes have been associated with the pathogeneses of several psychiatric disorders, including bipolar disorder and schizophrenia, as well as neurodegenerative disorders, such as Parkinson’s disease^42,43,44^. Phosphatidylinositol consists of a family of phospholipids, known as phosphoinositides, that play key roles in cell physiology and signaling^45^. Dysfunction of the phosphatidylinositol pathway has been implicated in and suggested as a therapeutic target for OCD^46,47^. Of note, phosphatidylinositol 4,5-bisphosphate is the main lipid binding partner of proteins involved in clathrin-dependent endocytosis^48^, suggesting that aberrant phosphatidylinositol binding may lead to the dysfunction of clathrin-dependent endocytosis processes in OCD pathogeneses. However, more research will be needed to validate the involvement of these two biological pathways in childhood-onset OCD. Nonetheless, this study provides the first evidence of their dysregulation as potential genetic risk factors for the disorder.

Our functional enrichment meta-analyses also revealed an overrepresentation of genes expressed in various brain tissues and cells^11,12,35,36,39^. Most interestingly, we found an enrichment of genes involved in the fetal brain^39^, suggesting that genes harbouring DNVs in childhood-onset OCD probands may also play a role in neurodevelopment. This is supported by the observed enrichment of genes expressed in astrocytes and oligodendrocytes^39^, two key cells in the mammalian brain with crucial roles in neurodevelopmental processes^49,50^. Also of note, we observed an overrepresentation of genes expressed in the cingulate gyrus and the prefrontal cortex^39^, two key components of the cortico-striatal-thalamo-cortical (CSTC) system^51^. The CSTC neural loop is involved in many cognitive and emotional processes, including reward-based learning, decision making, and goal-directed behaviour^51^. It is also involved in important motor functions, including procedural and habit learning, the selection and execution of appropriate actions, action inhibition, and impulsivity control^51^. The CSTC has been consistently implicated in obsessive-compulsive tendencies and its dysfunction is widely hypothesized to underlie OCD^1,51^. Altogether, our findings support two ongoing hypotheses: that childhood-onset OCD is a potential neurodevelopmental subtype of the disorder^52^, and that the CTSC system is a key neural loop involved in the pathogenesis of OCD^1,51^.

There exist several potential limitations to the present study. Our sample size is small for a DNV analysis, limiting our statistical power to detect exome-wide significant genes as well as statistically significant associations within our regression models. Most notably, the linear regression models investigating the effect of advanced parental age at proband conception were likely underpowered due to our small sample size given it is widely recognized that advanced paternal age is associated with an increased *de novo* mutation rate^53^. Our small sample size also calls for follow-up analyses with larger cohorts to confirm and clarify our findings. Moreover, many of our probands have psychiatric comorbidities (including tic, anxiety, eating, depressive, social, and language disorders). Consequently, some of the genes identified in this study may represent those contributing to multiple comorbid psychiatric disorders or to the general risk for psychopathologies rather than childhood-onset OCD specifically. Furthermore, without access to age- and ancestry-matched control trios, an enrichment analysis of dnSNVs in cases compared to controls could not be performed for our cohort to support the role of dnSNVs in childhood-onset OCD for French Canadians. The use of self-reported ancestry may also represent a limitation as self-reports can be inaccurate for determining genetic ancestry as they are often influenced by non-genetic information^54^. While QC steps were implemented to ensure a genetically homogenous cohort indicative of French-Canadian genetic ancestry, it remains possible that some of the trios included in the analyses are of broader European genetic ancestry. Additionally, by virtue of using WES, we were limited to the detection of dnSNVs located in the sequenced exome of our probands. As such, we were unable to detect dnSNVs located in non-coding regions that may be contributing to childhood-onset OCD risk. Finally, as in any DNV study, the possibility that there may be undetected dnSNVs in our available proband data relevant to our phenotype of interest also cannot be excluded.

Nonetheless, this study presents a framework for conducting DNV analyses of complex psychiatric disorders in genetically isolated populations wherein sample sizes are often limited. Using this framework, we provided the first list of candidate childhood-onset OCD genes that are specific to the French-Canadian population. We also identified biological mechanisms that may be underlying childhood-onset OCD risk and provided further support for a shared underlying susceptibility to psychiatric disorders. While larger sample sizes with deep phenotypic information will be needed to elucidate the role of dnSNVs in childhood-onset OCD symptom severity, we have provided a simple statistical model for investigating this relationship and emphasize the value of robust clinical data collection for these models. Altogether, this study sets the foundation for potential future studies with larger sample sizes that could afford greater resolution for detection via increased power of discovery. Future analyses should also focus on French-Canadian adult-onset OCD cases to better understand the role of dnSNVs in this patient subgroup.

## Data Availability

All dnSNVs identified in this study are listed in Table 1. Cohort descriptives for the probands and their unaffected parents that were sequenced as part of this study are available in Supplementary Table 1. Complete information on QC and DNV filtration preformed throughout this study are described in Supplementary Table 2.

QC was performed using GATK v3.8 (https://gatk.broadinstitute.org/hc/en-us) and PLINK 2.0 (https://www.cog-genomics.org/plink/2.0/). Blacklist variants were excluded using the ENCODE Blacklist Regions GRCh37 v2 (https://github.com/Boyle-Lab/Blacklist) and bedtools v2.30.0 (https://bedtools.readthedocs.io/en/latest/index.html). Kinship analysis and PCA were performed using SNPRelate v1.24.0 (https://github.com/zhengxwen/SNPRelate). PCA also used tidyverse v1.3.2 (https://www.tidyverse.org/) and the 1000 Genomes Reference Panel GRCh37 Phase 3 v5a (https://www.internationalgenome.org/). DNV calls were annotated using Ensembl VEP v107 (https://www.ensembl.org/info/docs/tools/vep/script/vep_download.html). The deleteriousness of the dnSNVs was scored using CADD GRCh37 v1.6 (https://cadd.gs.washington.edu/). MAFs of the dnSNVs were determined using gnomAD v2.1 non-neuro (http://www.gnomad-sg.org/downloads/). IGV v2.13.0 was used to visually inspect the dnSNVs (https://github.com/igvteam/igv). The enrichment of dnSNVs by variant class was performed using denovolyzeR v0.2.0 (https://github.com/jamesware/denovolyzeR). GTEx v8 was used to determine the expression levels of candidate genes in brain tissues (https://gtexportal.org/home/). EnrichR v3.2 (https://maayanlab.cloud/Enrichr/) and g:Profiler g:GOSt v0.2.3 (https://biit.cs.ut.ee/gprofiler/gost) were used for the functional enrichment meta-analyses.

## Acknowledgements

We are grateful to all the families who took part in this study. The authors thank Vessela Zaharieva for assistance in clinical coordination; Daniel Rochefort and Helene Catoire for assistance in sample management; and Jasmine Al-Shami for assistance in digitizing patient clinical data.

## Author Information

Conceptualization: K.B., J.P.R., Z.S., P.A.D., G.A.R.; Data Curation: D.S., B.B., J.M., G.L., G.A.R.; Formal Analysis: K.B., J.P.R.; Funding Acquisition: P.A.D., G.A.R.; Investigation: K.B.; Methodology: K.B., J.P.R., Z.S., M.M., D.S.; Project Administration: K.B., P.A.D., G.A.R.; Resources: D.S., P.A.D., G.A.R.; Supervision: P.A.D., G.A.R.; Validation: K.B.; Visualization: K.B.; Writing – Original Draft: K.B.; Writing – Review & Editing: K.B., J.P.R., Z.S., M.M., P.A.D., G.A.R.

## Ethics Declaration

Written informed consent was obtained for all individuals or their legal representatives. All data were deidentified. Ethical approval was obtained from the McGill University Health Centre (MUHC) Centre for Applied Ethics Research Ethics Board (REB) Neurosciences-Psychiatry (NEUPSY) panel (IRB00010120).

