## Supplementary Figures for "*De novo* variant analysis of childhood-onset obsessive-compulsive disorder in the French-Canadian population"

### Supplemental Information

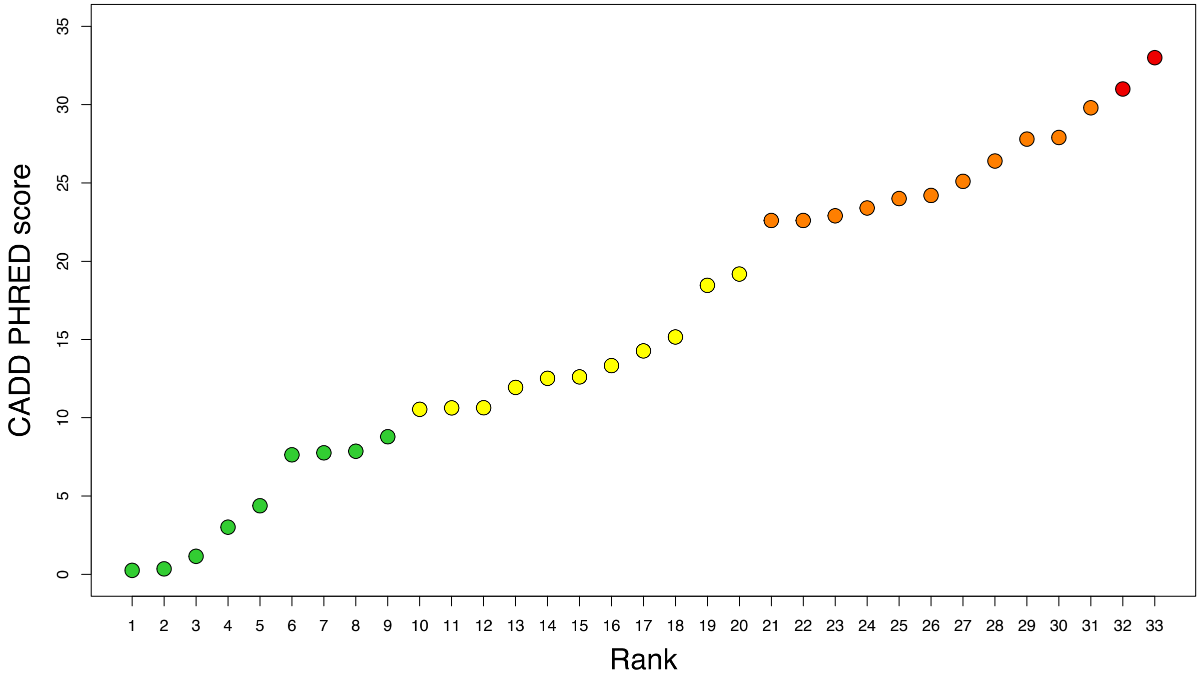


**Supplementary Figure 1. Distribution of CADD scores of the 34 dnSNVs identified in the 36 French-Canadian childhood-onset OCD probands.** Each point represents one dnSNV. dnSNVs are ranked in ascending order of their CADD scores. The 10 dnSNVs with CADD scores below 10 are in green. These variants are below the threshold of pathogenicity according to the CADD score system. The 11 dnSNVs with CADD scores between 10-20 are in yellow. These variants are predicted to be in the top 10% of deleterious substitutions in the human genome. The 11 dnSNVs with CADD scores between 20-30 are in orange. These variants are predicted to be in the top 1% of deleterious substitutions in the human genome. The two dnSNVs with CADD scores above 30 are in red. These variants are predicted to be in the top 0.1% of deleterious substitutions in the human genome.

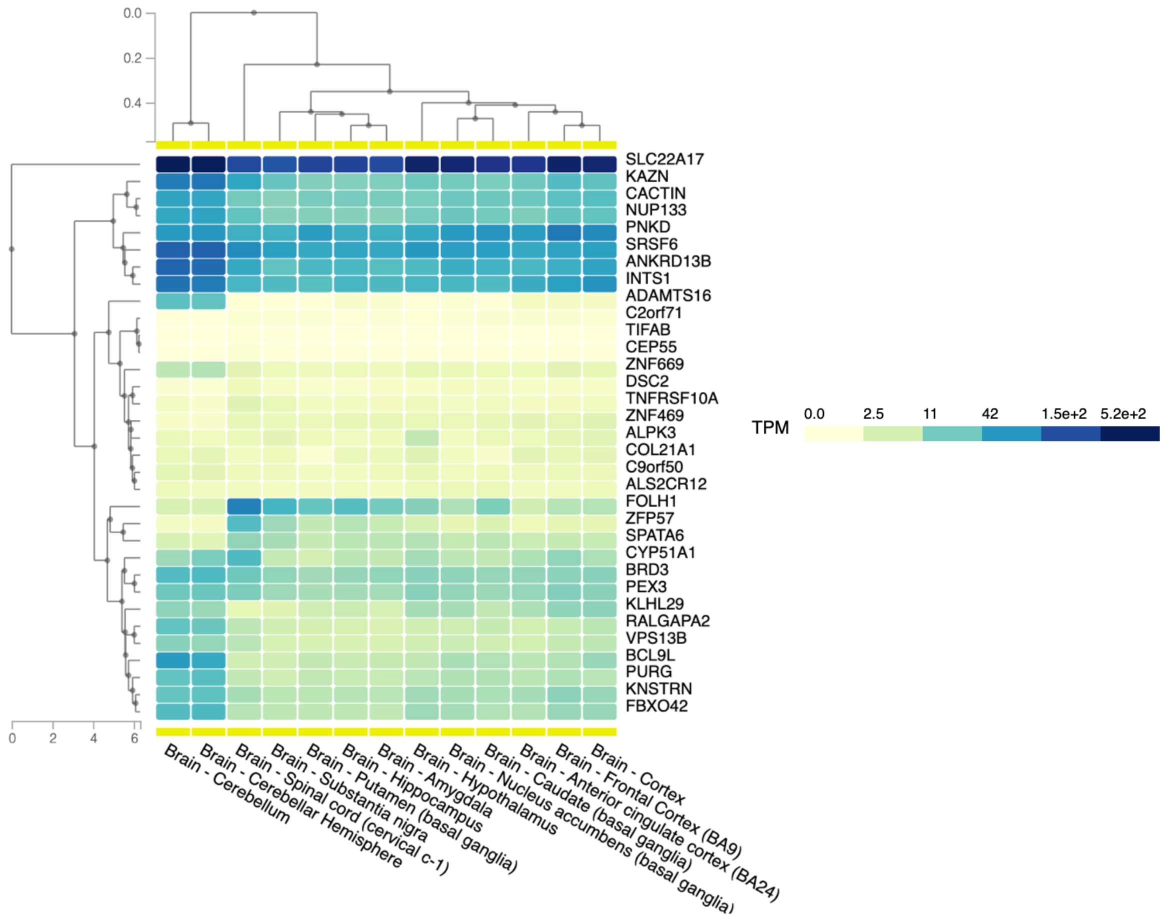


**Supplementary Figure 2. Expression of genes harbouring dnSNVs in French-Canadian childhood-onset OCD probands in brain tissues from the GTEx Project.** Tissue-specific expression levels are reported in transcripts per million (TPM). All 34 genes were expressed in at least one or more brain tissues (i.e., TPM > 0.0). Brain tissues were clustered based on the similarity of their gene expression profiles. Genes were clustered based on the similarity of their expression levels across the selected brain tissues.
